# A prevalence-incidence mixture model for interval-censored screening and post-treatment surveillance data in a population with a temporarily increased disease risk

**DOI:** 10.1101/2025.04.19.25325991

**Authors:** Kelsi R. Kroon, Johannes A. Bogaards, Johannes Berkhof

## Abstract

To optimize screening and surveillance programs, it is essential to have information on the duration between the onset of a high-risk condition and treatable disease. Modelling of this duration is complicated when the high-risk condition only applies to some of the subjects and/or is temporary (e.g., oncogenic infections). Additionally, interval-censored data between visits makes exact disease onset time unknown and some of the disease cases may already be present at baseline. We propose a prevalence-incidence mixture model for interval-censored data to estimate cumulative disease risk based on individual risk factors and to estimate the mean duration of the high-risk condition. Subjects without the high-risk condition may also develop disease which is modelled by adding background risk. Parameters are estimated with an Expectation-Maximisation (EM) algorithm with weakly informative Cauchy priors. The duration between the baseline high-risk condition and treatable disease is directly obtained from the model. A score test is derived to check for poor fit. The algorithm is validated through simulation studies and applied to Dutch cervical cancer screening and post-treatment surveillance data. The screening example is longitudinal data of women who tested positive for human papillomavirus (HPV) at baseline. Some of these had undiagnosed, treatable disease (CIN2+) at baseline, some developed CIN2+ during follow-up, and some were assumed to clear the infection. The second example is surveillance data of women treated for CIN2+. They either had residual CIN2+ at baseline, were HPV-infected (without CIN2+) despite the treatment or were HPV-uninfected. The HPV and disease status at baseline were not observed. Our EM algorithm showed very good convergence for simulated data and the parameter estimates were close to the true values for realistic sample sizes. In the examples, our model had lower Akaike’s Information Criterion values than models with Weibull, B-spline and lognormal hazards for incident events (difference 0.7 to 26.7). To conclude, our prevalence-incidence mixture model can provide an accurate fit to the data and can be used for designing risk-based screening and surveillance programs.

## 1. Introduction

In public health and medical research, prediction models play a crucial role in identifying individuals who are at highest risk of developing a disease. In screening and post-treatment surveillance, there are two potential scenarios for disease detection. Individuals who are screen-positive or treated for disease may have underlying *prevalent* disease at baseline, or disease might develop in-between two visits (*interval-censored incident disease*). Furthermore, incident disease can develop either from the baseline condition or from a newly acquired condition during follow-up. The baseline condition may only apply to a proportion of subjects and may be temporary so that the cumulative disease risk is likely to level off.

A motivating example is human papillomavirus (HPV) infections, which cause almost all cervical cancers worldwide^1^ and are also associated with several other cancers and warts^2^. After an initial HPV infection, individuals are at increased risk of developing a high-grade cervical lesion (CIN2+) until the infection clears. To optimize a cervical cancer screening or post-treatment surveillance program, it is essential to estimate the duration between the high-risk condition and CIN2+.

Non-parametric and parametric models have widely been used for estimating the cumulative risk but they have limitations when applied to our setting. Non-parametric cumulative incidence models^3–6^ assume that there are no prevalent disease cases. This can be accounted for by defining a very small interval with a left bound of zero^7^. However, a non-parametric model does not have covariates which are crucial when designing risk-based screening or surveillance programs. Parametric models allow for more flexibility with respect to the modelling of the effects of covariates. Prevalence-incidence mixture models have been proposed^7,8^ which assume a logistic regression model for prevalent disease, some of which are undiagnosed at baseline, and a parametric model (e.g. Weibull) for incident disease. These models may provide an accurate fit of the incidence function, but do not provide credible estimates of the mean time to incident disease. This is because they assume that the cumulative incidence approaches 1 for large time *T* whereas the observed incidence is typically low in screening and post-treatment surveillance settings. A reason for the low incidence is that only a proportion of the subjects without prevalent disease at baseline have a high-risk baseline condition^9^. For example, in post-treatment surveillance studies, subjects are at high risk of a recurrent lesion when not all causative cells were removed by the treatment. Furthermore, the high-risk condition may be temporary, for example, because a competing event prevents progression from the high-risk condition to disease. If the competing event is observed during follow-up, then a prevalence-incidence competing risk model can be specified^10^. However, there are several situations where such a model cannot be applied. For example, in screening studies the competing state is not measured during follow-up when follow-up is done in accordance with national guidelines and the new screening instrument has not yet been implemented.

We propose to capture the heterogeneity in the data through a prevalence-incidence mixture model where a proportion of prevalent disease cases is undiagnosed at baseline and subjects without prevalent disease have a temporarily increased disease risk that converges with the background risk over time. Incident cases developed from the baseline condition are assumed to follow a mixture of an exponential distribution and non-progressive condition, whereas other incident disease cases are described by adding background risk. The model for incident disease developed from the baseline condition is mathematically equivalent to a model with an exponential time distribution for two competing events, of which one is not observable^11^. To examine whether the model is violated by the data, we present a score test comparing the exponential distribution for incident disease from the baseline condition against a Gamma alternative.

The article is structured as follows. In Section 2, we describe the model and an expectation-maximisation (EM) algorithm for estimating the model parameters with interval-censored screening data, which has been implemented in an R package. In Section 3, we present simulation studies to verify that the algorithm can accurately recover true parameter values and a score test to check the exponential distribution for incident disease from the baseline condition. In Section 4, we apply the model to estimate the cumulative risk of CIN2+ in two different data sets. The first dataset consists of cytological and histological follow-up of a screening population of women who tested positive for HPV at baseline. Some of these women had undiagnosed CIN2+ at baseline, some developed CIN2+ during follow-up and some were assumed to clear the infection. The second dataset consists of cytological and histological follow-up of a cohort of patients who were treated for CIN2+ at baseline. Some of these women had (undiagnosed) residual CIN2+ at baseline, some were assumed to be HPV-infected despite the treatment and were at high risk of developing recurrent CIN2+ during follow-up, and some were assumed to be HPV-uninfected at baseline. The HPV status at baseline was not measured. Finally, in Section 5 we conclude and discuss our findings.

## 2. Model

Due to the nature of screening programs and disease surveillance, the exact time of onset of disease is not observed. Instead, there is only an interval of the last time an individual with the baseline condition was observed without the disease, and the first time that the disease was detected. If the disease did not develop during follow-up, this interval will be right-censored. Further, some individuals are immediately referred at baseline, therefore it can be determined whether the disease is present at baseline. However, some individuals are not referred at baseline but the disease is detected at their first visit, in which case it is unknown whether the disease was present at baseline.

For individuals *i* = 1, …,*n* who have an elevated risk due to a baseline condition, let (*L*_*i*_, *R*_*i*_ ]be the interval when the disease develops, where *L*_*i*_ is the last time the *i* ^*th*^ individual was observed without disease and *R*_*i*_ is the first time that disease is detected for the *i* ^*th*^ individual. We allow *R*_*i*_ = ∞ to include individuals who do not develop disease during follow-up. Let *z*_*i*_ be an indicator variable for prevalent disease at baseline (where *z*_*i*_ = 1 means that the disease was present at baseline), which, as described above, is unknown for some individuals. Let ***θ***_1_, ***θ***_2_ be disjoint vectors of parameters, and let ***x***_1_ and ***x***_2_ be subject-specific vectors of covariates for prevalence and progression respectively. Let *π* (***x***_1_; ***θ***_1_) be the probability of having the disease at baseline (prevalent disease). Then the probability that the disease has developed before time *t*, i.e. the time to disease, denoted by *T*, lies in [0, *t*), is a mixture distribution given by

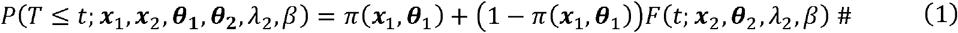

where *F*(*t*; ***x***_2_, ***θ***_2_, *λ*_2_, *β*) is the cumulative risk of incident disease. Among incident cases there are two independent causes, disease can either develop from the baseline condition or from another condition (e.g. a new infection acquired after baseline). Events from the baseline condition follow a competing events framework, where the baseline condition either progresses to the disease or transitions to an unobservable, competing state, both of which are assumed to follow an exponential distribution.

Let us assume a competing risk framework with progression parameter *λ*_1_ (***x***_2_, ***θ***_2_) and clearance parameter *λ*_2_. Then the cumulative risk of developing disease from the baseline condition at time *t* is given by

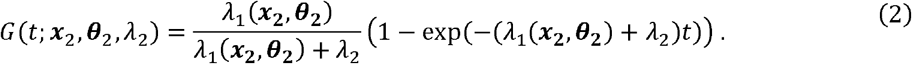

In this specification, δ(***x***_2_, ***θ***_2_) = *λ*_1_ (***x***_2_, ***θ***_2_)/(*λ*_1_ (***x***_2_, ***θ***_2_) + *λ*_2_) is the probability of disease during follow-up developed from the baseline high-risk condition. The expected time from the high-risk condition and the event is given by 1/(*λ*_1_ (***x***_2_, ***θ***_2_) + *λ*_2_). So far, we assumed that all subjects have a high-risk condition at baseline, but the model can also be applied when it is not known who is at high-risk of developing disease. In this situation, only a proportion of the subjects have a high-risk condition and either develop disease or transition to a competing, unobservable event. The expected duration from the high-risk condition to disease is still given by 1/(*λ*_1_ (***x***_2_, ***θ***_2_) + *λ*_2_), but δ(***x***_2_, ***θ***_2_) is now the proportion of subjects who have a high-risk condition and develop disease from this high-risk condition. The progression parameter *λ*_1_ (***x***_2_, ***θ***_2_) can in both situations be interpreted as the derivative of the cumulative incidence *G* (*t*; ***x***_2_, ***θ***_2_, *λ*_2_) at *t* = 0, i.e. *G*′ (0; ***x***_2_, ***θ***_2_, *λ*_2_).

For events that do not develop from the baseline condition, we define the cumulative background risk of developing disease, *H* (*t*; *β*), given by:

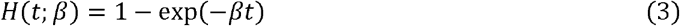

where *β* is the background hazard. Now, we get the following model for incident disease:

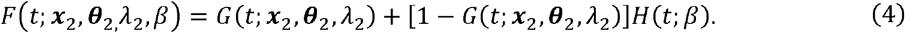

The model allows for the inclusion of baseline covariates for the prevalence parameter and the rate parameter *λ*_1_ (***x***_2_, ***θ***_2_). For the proportion of prevalent disease, *π* (***x***_1_, ***θ***_1_), a logistic regression model is used to include covariates:

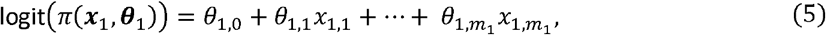

where *m*_1_ is the number of covariates included in the logistic model for prevalent disease. For both *λ*_1_ and *λ*_2_, a log transformation is used in which case the covariates for *λ*_1_ have a proportional effect on its hazard:

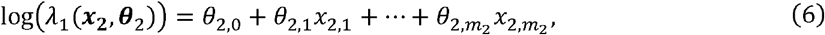

where *m*_2_ is the number of covariates used for the rate parameter *λ*_1_.

In principle, the background risk *β* can also be linked to covariates, but this is not considered here as its regression coefficients tend to be less well identified as those of *π* and *λ*_1_. The main role of *β* is to describe how the baseline hazard is influenced by the risk of an event that did not develop from the baseline condition.

In our model, the regression coefficient *θ*_2,*i*_ (*i* ≥ 1) can also be interpreted as the change in the log-odds of progression probability δ (***x***_2_, ***θ***_2_) associated with a one-unit increase in the corresponding covariate. To see this, let 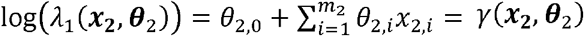, and log(*λ*_2_)= *ϕ*. Then:

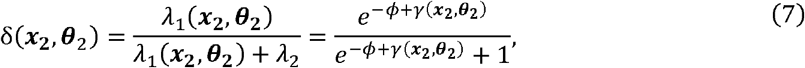

so the logit is linear in the covariates: logit δ (***x***_2_, ***θ***_2_)= −*ϕ* + *γ* (***x***_2_, ***θ***_2_).

A variant of the traditional expectation-maximisation (EM) algorithm called the EM gradient algorithm^12^ is used to estimate the parameters. The EM gradient algorithm approximately solves the maximisation step of the EM algorithm by using a single Newton step on the expected log-likelihood instead of directly solving the maximisation, and it has been shown to have similar convergence properties as the standard EM algorithm^13^. Details about the estimation can be found in the Supplementary Appendix.

A situation might arise where there are no or a very low number of subjects with a specific combination of a covariate and the outcome. This causes identifiability issues when estimating the parameter for this covariate, and although the algorithm may converge, it is likely that the confidence intervals for the parameter will be very large. To address these issues in the logistic prevalence mixture, we choose a Cauchy distribution with a centre of 0 and scale of 2.5, as recommended by Gelman et al. (2008)^14^, as a weakly informative prior. The first and second derivatives of the log of the Cauchy (0, 2.5) distribution with respect to each parameter can be directly evaluated in the maximisation step and added to calculations of the gradient and Hessian, respectively. Credible intervals for the parameters in the model with a Cauchy prior can be obtained through Taylor expansion^15^.

## 3. Simulation study

### 3.1 Validation of model fit

#### Simulation study I: setup

We assessed the performance of our model in a simulation study where data was simulated according to the following settings. For the disease process, values for the parameters ***θ***_1_, ***θ***_2_, *λ*_2_ and *β* were set, and for each subject *i*, a Bernoulli random variable for prevalent disease, *P*_*i*_, was drawn with probability *π*(***x***_1_, ***θ***_1_). In subjects without prevalent disease (i.e. *P*_*i*_ = 0), a second Bernoulli variable for incident disease from the baseline condition, *A*_*i*_, was drawn with probability δ(***x***_2_, ***θ***_2_) . Additionally, two event times were generated for incident disease developed from the baseline condition (*T*_1_) and from the new condition acquired after baseline (*T*_2_). *T*_1_ was drawn from an exponential distribution with rate parameter *λ*_1_ (***x***_2_, ***θ***_2_) + *λ*_2_ if *A*_*i*_ = 1, and set to infinity otherwise and *T*_2_ was drawn from an exponential distribution with rate parameter *β*. We then took the minimum of *T*_1_ and *T*_2_ as the onset time of incident disease, i.e., *T*_D_ = min(*T*_1_, *T*_2_).

Screening (or post-treatment surveillance) times were drawn from a Beta(20, 20) distribution scaled to have a mean around 3-yearly intervals until there was at least 25 years of follow-up. Attendance of each individual at each screening visit was drawn from a Bernoulli distribution. This resulted in left and right intervals representing the last time an individual was observed before disease onset and the first time after disease onset.

The following parameter values for prevalence, progression, and clearance were used for the simulation study without covariates: *π* = log (0.20), log (*λ*_1_)= −1, and log (*λ*_2_)= −1.5. These parameter values were loosely based on values observed in cervical cancer screening studies. We investigated the effect of simulation settings on model performance by varying the (1) sample size (N=1000 or 10,000), (2) attendance probability per screening round (0.75 or 1), (3) value of background risk (*β* = 0, 0.005, 0.01, or 0.02) and (4) weakly informative prior (none or Cauchy(0,2.5)). This resulted in 32 separate simulation conditions.

The simulation study was repeated with baseline covariates, again loosely based on values in cervical cancer screening studies. A continuous variable Age was generated from a Uniform(30, 70) distribution to reflect age at baseline, and a binary variable HPV16 (e.g., positive for HPV genotype 16 at baseline) was generated with probability of being equal to 1 set at a 0.3. This parameter was assumed to positively affect both the prevalence and progression probabilities in the model. The following model was used:

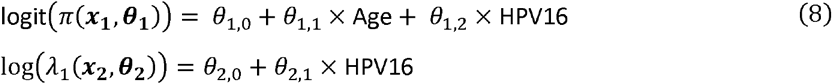

with parameters set at: ***θ***_**1**_ = { *θ*_1,0_ = −3, *θ*_1,1_ = 1, *θ*_1,2_ = 1}, ***θ*_2_** = {*θ*_2,0_ = −1, *θ*_2,1_ = 2}, and *λ*_3_ = −1.5. All simulated data sets were fit with our EM algorithm using 10 different random starting values per data set. For each simulation setting 200 data sets were generated and the median estimate, the percent bias, and 95% confidence/credible interval coverage probability (CP) were calculated (shown in Figure 1). Additionally, the time that it takes for the algorithm to converge and the convergence rate were calculated for models with and without covariates in the setting with low background risk (*β* = 0.005), imperfect attendance probability (0.75) and priors included.

**Figure 1.**
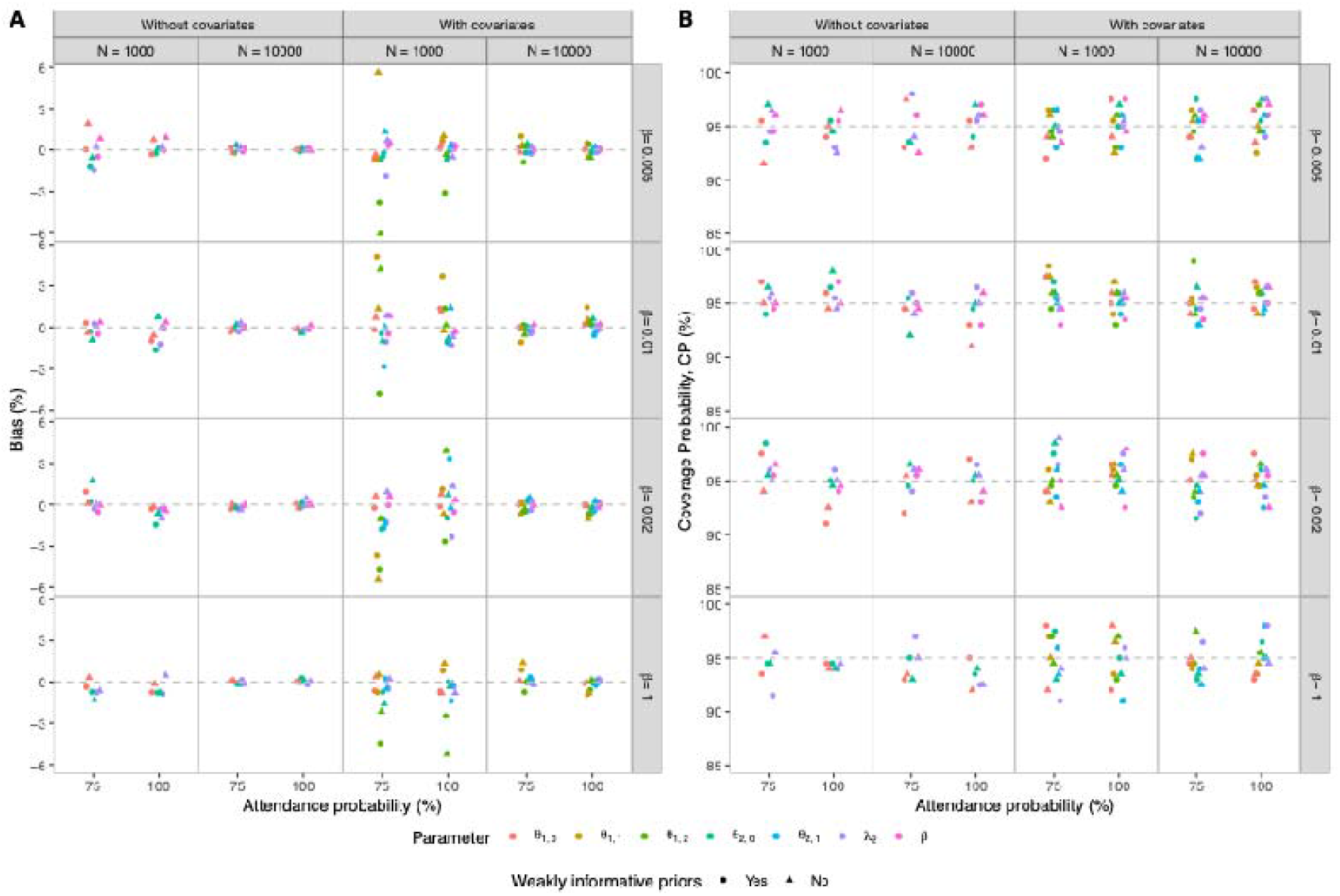
Simulation study results without covariates and with covariates for combinations of prior inclusion, sample size (N), attendance probability and value of background risk. Panels A and B show bias and coverage probability, respectively. Without covariates, parameters are prevalence (*θ*_1,1_), progression (*θ*_2,1_), clearance (*λ*_2_), and background risk (*β*). With covariates, parameters are prevalence intercept (*θ*_1,0_), effect of age on prevalence (*θ*_1,1_), and effect of HPV16 on prevalence (*θ*_1,2_), progression intercept (*θ*_2,0_), effect of HPV16 on progression (*θ*_2,1_), clearance (*λ*_2_), and background risk (*β*).

#### Simulation study I: results

The model performed well in percentage bias and coverage across all parameters in simulations, regardless of covariates, attendance probability, or priors, for low background risk (Figure 1). Without covariates, percentage bias ranged from -1.64% to 1.93% (N=1000) and -0.42% to 0.45% (N=10,000), with coverage probability between 91.0% to 98.5% (N=1000) and 91.0% to 98.0% (N=10,000). Percentage bias ranged between -1.45% and 1.93% at attendance probability of 0.75 and between -1.64% and 0.91% at perfect attendance. Percentage bias ranged from -1.30% to 1.93% without priors and from -1.64% to 0.95% with weakly informative priors. Attendance probability and inclusion of priors had minimal impact on coverage probability. With covariates, similar trends were observed. For N=1000, percentage bias ranged from -6.05% to 5.63%, with coverage probability between 91.0% to 99.0%. For N=10,000, the absolute percentage bias was lower than for N=1000 (<1.45% across settings), and the coverage probability ranged from 91.5% to 99.0%.

The average CPU time (in seconds) to fit each of the 200 simulated data sets increased with covariates. Without covariates, average time was 2.42s (N=1000) and 19.77s (N=10,000), and with covariates the average time rose to 14.96s (N=1000) and 143.85s (N=10,000). There were no cases of non-convergence in any of the settings.

### 3.2 Checking exponential progression rate

#### Score test

The exponential progression rate for events developed from the baseline condition may be violated by the data even after adjusting for competing, unobserved events and events that did not develop from the baseline condition. This limits the applicability of the model when used to support decision making for a screening or post-treatment surveillance protocol with multiple test moments. An example is the outcome cancer developing through multiple stages which can be described by assuming an increasing hazard since onset of the baseline condition. We evaluated the fit of an exponential time distribution for events developed from the baseline condition by devising a score test with a Gamma distribution as alternative. If the progression to disease follows a Gamma distribution with shape parameter *k* and rate parameter *λ*_1_, then the cumulative incidence function for events developed from the baseline condition becomes

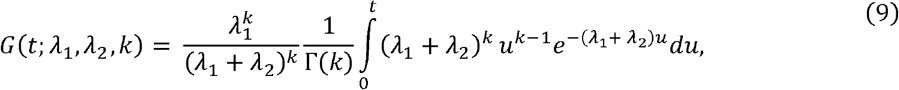

which in turn is the distribution function of a Gamma variable with shape parameter *k* and rate parameter *λ*_1_ + *λ*_2_ multiplied by the constant 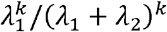.

The null hypothesis is that the Gamma shape parameter, *k*, in equation (9) equals one, thereby reducing the Gamma distribution to an exponential distribution. To calculate the gradient and Hessian of the log-likelihood function evaluated at *k* = 1, we used numerical methods to obtain the first and second derivatives of *G* (*t*) in equation (9) with respect to the shape parameter. Let 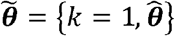 be the constrained maximum likelihood estimator (MLE) where 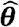 is the MLE for the remaining parameters, 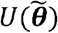 is the score vector, and 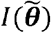 is the Fisher information matrix. Then the score test statistic is given by 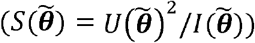 which is asymptotically Chi-squared distributed with 1 degree of freedom under the null.

#### Simulation study II: setup

Simulation studies were performed to evaluate the power and Type 1 error of the model under varying combinations of the (1) shape parameter, (2) background risk, and (3) sample size. Data was generated according to the procedure described previously, however in this case the Bernoulli variable for incident disease developed from the baseline condition, *A*_*i*_, was drawn with probability 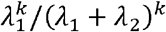 and *T*_1_ was generated from a gamma distribution with shape parameter *k* and rate parameter *λ*_1_ + *λ*_2_, according to equation (9). The shape parameter was set at either *k* = 1, 1.5 or 2, where *k* = 1.5 and *k* = 2 are time distributions with increasing hazards. Background risk was set at either *β* = 0, 0.005, 0.01, or 0.02, and was assumed to be either known and fixed at its correct value or estimated with the EM algorithm. Sample size was either *N* = 1000 or 10,000. This makes 2 (*N*) × 3 (*k*) × 3 (*β*) = 36 combinations in case the background risk is positive, plus 2 (*N*) × 3 (*k*) = 6 combinations in case the background risk is zero resulting in 42 combinations. Data sets were generated for each combination until 100 data sets had a successful model fit.

#### Simulation study II: results

[Place Table 1 near here] The results are in Table 1. For N=1000, the Type I error ranges from 0.03 to 0.10 and for N=10,000, it does not exceed 0.05. For N=1000, the power under *k* > 1 ranges from 0.43 to 0.99 when *k* = 2 and is negligible or low when *k* < 2. For N=10,000, the power ranges from 0.71 to 1.00.

**Table 1.**
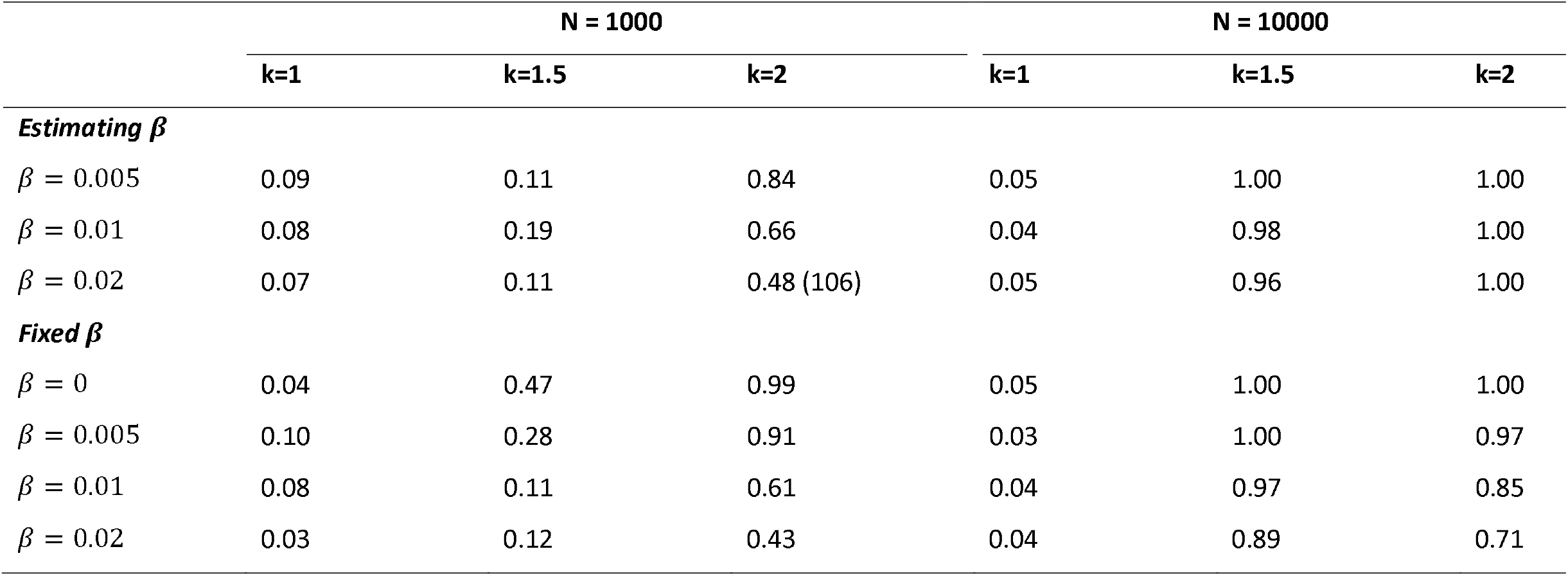
Power of the Gamma score test for different combinations of background risk (*β*), sample size (N) and shape parameter (*k*). Data sets were generated for each combination until 100 data sets had a successful model fit. The brackets show the number of data sets that were required to have 100 converged model fits for each simulation setting.

## 4. Examples

### 4.1 Cervical cancer screening data

The Population-based screening study Amsterdam (POBASCAM) is a randomized controlled screening trial (Netherlands trial registration ID: NTR218) in the Netherlands consisting of women from the Dutch population recruited between 1999 and 2002, and the maximum follow-up was 19 years. The study has been described before^16,17^. In this application, we used a subset of 2,269 women who were HPV-positive at their first screening date (baseline). The date of the first histology test reporting CIN2+ was set as the right bound. If an index smear was reported within 3 months before the first CIN2+ histology diagnosis, it was used as the right bound instead. For the left bound, the most recent normal cytological finding and/or histological finding reporting <CIN2 occurring before the right bound was used.

The information loss, measured by the AIC, was low for our model without covariates (AIC = 4740.12) compared to the logistic-Bspline model^7^ with five knots (AIC = 4740.83), the logistic-Weibull (AIC = 4767.84) and the logistic-lognormal model (AIC = 4747.46).

[Place Table 2 near here] Table 2 gives parameter estimates with 95% confidence intervals, log-likelihood values and score test results for our model with abnormal baseline cytology defined as a binary variable in the prevalent disease regression equation, and HPV16 genotype defined as a binary variable in the prevalent and incident disease regression equations. The selection of the variables was based on HPV literature, which reported cytology as prognostic for prevalent disease and HPV16 genotype as prognostic for prevalent and future disease^18,19^. The score test was not significant for the models with and without background risk. Adding background risk improved the fit of the model (LR 5.56, p-value 0.018), and adding the Cauchy prior reduced the width of the credible intervals in the model with background risk.

**Table 2.** Parameter estimates (95% confidence/credible intervals (CI/ CrI)) for the POBASCAM study application with and without the Cauchy prior.

|  |  |  | Without priors or<br>background risk | Without priors, with<br>background risk | With priors, with<br>background risk |
| --- | --- | --- | --- | --- | --- |
| Parameter |  |  | Estimate (95% CI) | Estimate (95% CI) | Estimate (95% CrI) |
| Prevalence | Intercept | $\hat{\theta}_{1,0}$ | -5.28 (-6.84, -3.73) | -12.83 (-519.75, 494.09) | -5.66 (-7.73, -3.59) |
| | Effect of HPV16 | $\hat{\theta}_{1,1}$ | 1.22 (0.9, 1.54) | 1.22 (0.89, 1.54) | 1.21 (0.88, 1.53) |
| | Effect of abnormal cytology | $\hat{\theta}_{1,3}$ | 4.55 (2.99, 6.11) | 12.09 (-494.83, 519.02) | 4.93 (2.86, 7) |
| Progression | Intercept | $\hat{\theta}_{2,0}$ | -3.4 (-3.59, -3.21) | -3.47 (-3.73, -3.21) | -3.5 (-3.78, -3.23) |
| | Effect of HPV16 | $\hat{\theta}_{2,1}$ | 0.96 (0.74, 1.18) | 1.23 (0.89, 1.56) | 1.21 (0.87, 1.55) |
| Clearance | | $\hat{\lambda}_2$ | -2.05 (-2.25, -1.85) | -1.58 (-1.97, -1.2) | -1.61 (-2.01, -1.21) |
| Log of background risk | | $\log(\hat{\beta})$ | | -- | -5.23 (-5.98, -4.49) |
| Log-likelihood |  |  | -2037.36 | -2034.58 | -- |
| Gamma score test p-value |  |  | 0.97 | 0.96 | 0.49 |

HPV16: human papillomavirus genotype 16

For HPV16-positive women, the mean time to CIN2+ was 3.32 years (95% CI: 2.23, 4.43) and the proportion of women who developed CIN2+ from the baseline infection was 0.33 (95% CI: 0.29, 0.38). For HPV16-negative women, the mean time to CIN2+ was 4.34 years (95% CI: 2.83, 5.86) and the proportion of women who developed CIN2+ from the baseline condition was 0.13 (95% CI: 0.10, 0.16).

### 4.2 Cervical post-treatment follow-up data

After treatment of CIN2+, there is a chance of residual disease due to incomplete excision (prevalent disease) or HPV infected cells (the baseline condition) regrow to CIN2+. Women also remain at risk of developing a new CIN2+ (background risk)^20^. We used data from a long-term multi-cohort study following women after treatment for CIN2+ (Netherlands trial registration ID: NTR1468)^21^. This cohort study included 435 women who were monitored with HPV testing and cytology at 6, 12, and 24 months after treatment. Thereafter, women were followed through the routine screening programme with 5-year intervals. The maximum follow-up was 21 years.

We compared our model without covariates to other prevalence-incidence models using Akaike’s Information Criterion (AIC). Our model (AIC = 610.13) fitted slightly better than a logistic-Bspline model with five knots (AIC = 617.39), logistic-Weibull (AIC = 614.64) and logistic-lognormal model (AIC = 613.48).

[Place Table 3 near here] Table 3 gives parameter estimates with 95% confidence intervals, log-likelihood values and score test results for three models with severity of disease at treatment as covariate (CIN3: 1 if CIN3+, 0 if CIN2) in the prevalent and incident disease regression equations. The score test was significant for a model without background risk (p-value = 0.0047), but was not significant when background risk was included (p-value = 0.51) indicating that an exponential progression model for events developed from the baseline condition and background risk for other events was not violated by the data. Adding background risk also significantly improved model fit (LR 8.52, p-value: 0.0035), but led to wide confidence intervals for the intercept and effect of CIN3. Smaller credible intervals could be obtained by adding a Cauchy prior.

**Table 3.** Parameter estimates (95% confidence/credible intervals (CI/ CrI)) for the post-treatment surveillance data application with and without the Cauchy prior.

|  |  |  | Without priors or<br>background risk | Without priors,<br>with background risk | With priors, with<br>background risk |
| --- | --- | --- | --- | --- | --- |
| Parameter |  |  | Estimate (95% CI) | Estimate (95% CI) | Estimate (95% CrI) |
| Prevalence | Intercept | $\hat{\theta}_{1,0}$ | -3.60 (-5.35, -1.85) | -4.33 (-8.7, 0.04) | -3.6 (-5.21, -1.99) |
| | Effect of CIN3 <sup>a</sup> | $\hat{\theta}_{1,1}$ | 1.15 (-0.66, 2.95) | 1.39 (-2.96, 5.74) | 0.62 (-1.05, 2.29) |
| Progression | Intercept | $\hat{\theta}_{2,0}$ | -3.41 (-4.31, -2.51) | -2.73 (-4.04, -1.42) | -2.73 (-3.8, -1.65) |
| | Effect of CIN3 <sup>a</sup> | $\hat{\theta}_{2,1}$ | 0.34 (-0.57, 1.25) | 0.58 (-0.67, 1.83) | 0.60 (-0.42, 1.62) |
| Clearance | Intercept | $\hat{\lambda}_2$ | -1.24 (-1.69, -0.78) | -0.20 (-0.77, 0.36) | -0.18 (-0.73, 0.37) |
| Log of background risk | | $\log(\hat{\beta})$ | -- | -5.76 (-6.61, -4.91) | -5.71 (-6.53, -4.88) |
| Log-likelihood |  |  | -303.13 | -298.88 | -- |
| Gamma score test p-value |  |  | 0.0047 | 0.51 | 0.24 |

<sup>a</sup> Histology variable is original baseline diagnosis (CIN2 = 0, CIN3 = 1).
CIN2/3: cervical intraepithelial neoplasia grade 2/3

For women with CIN3 at treatment, the expected time to recurrence was 1.05 years (95% CI: 0.45, 1.64) and the proportion that developed disease was 0.13 (95% CI: 0.04, 0.21). For women with CIN2 at treatment, the expected time to recurrence was 1.11 years (95% CI: 0.50, 1.72) and the proportion that developed disease was 0.07 (95% CI: 0.00, 0.14).

## 5. Discussion

We present a model for a population with a heterogenous disease risk that adds to the previously introduced concept of prevalence-incidence mixture models for the cumulative risk of disease^7,8^. By combining concepts from this model with additional parameters, we were able to decompose a complex process into simpler key components that together provide accurate estimates of the cumulative disease risk compared to non-parametric estimates.

To model progression, a competing risks framework with exponential distribution for disease progression was used which produced cumulative incidence curves with an AIC fit comparable to or lower than those of existing methods (i.e., Weibull, lognormal, B-splines).

A specific feature of our model is that it separates events that are caused by the baseline condition and events that are not caused by the baseline condition. Models that do not separate these events are less suitable when the aim is to estimate the duration from the baseline condition and treatable disease.

Our estimated model parameters can be used in Markov simulation models that are widely used to estimate the health gains and harms (i.e., QALYs) of new screening algorithms over a patient’s screening lifetime. Our score test can be used to detect lack of fit and hence be used to evaluate whether a Markov simulation model is violated by the data. If the model is consistent with the data, our model can be used to predict the impact of multiple rounds of screening and inform changes in the screening interval when moving towards a more personalised approach based on individual risk factors. Furthermore, our model can be used in post-treatment surveillance settings to assess how long the disease risk exceeds the background risk, warranting surveillance.

A limitation of our model is that it does not account for the sensitivity of the screening test results collected after baseline. This is important when determining the left and right bound of the interval-censored data. The bias in the cumulative incidence is expected to be small when the sensitivity is above 0.50, but can be substantial if the screening test sensitivity is lower^3^. Notably, the sensitivity of repeat cytology for detecting CIN2+ is about 0.85^22^.

Additionally, our model, when viewed as a competing risk model, does not use follow-up measurements from the baseline condition in order to assess clearance. Our model was motivated by cohort studies where the baseline condition is not reassessed at follow-up measurements. A Weibull prevalence-incidence mixture model with a clearance event was developed by Hyun (2020)^10^. This model still assumes that the high-risk condition can be measured without error and a possible path for future research is to account for the limited sensitivity of the screening test for assessing the high-risk condition during follow-up. A Bayesian prevalence-incidence mixture model was recently proposed that takes misclassification into account due to limited sensitivity of the screening test^23^. Such an approach, adapted to a competing risk setting, could be beneficial when analysing large data with detailed information recorded at every visit.

In conclusion, our model provides a flexible and interpretable approach for estimating disease risk that can be used to inform changes in the screening program and the incorporation of individual risk factors as we move towards a more personalized approach. Future work could explore incorporating test sensitivity of follow-up screening test results into the model.

## Data Availability

The data that support the findings of this study are available from the corresponding author upon reasonable request. The code for the accompanying R package is publicly available on GitHub (https://github.com/kelsikroon/PI3M).

## Acknowledgements

The authors thank C JLM Meijer for providing the POBASCAM data.

## Notes

**Funding:** This work was supported by the Horizon 2020 research and innovation program of the European Commission (RISCC project, grant agreement No 847845) (to KRK, JAB, and JB). The funders had no role in study design, data collection and analysis, decision to publish, or preparation of the manuscript.

**Conflict of interest:** KRK, JAB, and JB declare no conflicts of interests.

### Competing Interest Statement

The authors have declared no competing interest.

### Funding Statement

This work was supported by the Horizon 2020 research and innovation program of the European Commission [RISCC project, grant agreement No 847845] (to KRK, JAB, and JB). The funders had no role in study design, data collection and analysis, decision to publish, or preparation of the manuscript.

### Author Declarations

The POBASCAM trial (Trial registration ID: NTR218) was approved by the Medical Ethics Committee of the Vrije Universiteit (VU) University Medical Centre (Amsterdam, The Netherlands; no. 96/103) and the Ministry of Public Health (The Hague, The Netherlands; VWS no. 328650). All participants provided written informed consent. The post-treatment cohort study (Trial registration ID: NTR1468) was approved by the ethics board at all hospitals. All women provided written informed consent.

### Summary of Updates

Text throughout the manuscript has been updated for clarity of the model description and applicability.

