## Supplementary Appendix for "A prevalence-incidence mixture model for interval-censored screening and post-treatment surveillance data in a population with a temporarily increased disease risk"

**Supplementary Materials and Methods**

**1. Estimation with the Expectation-Maximisation Algorithm**

**1.1 Likelihood**

To construct the likelihood, subjects are divided into sets with missing and observed $z_{i}.$ Subjects with $z_{i}=1$contribute $\log\left( \pi\left( \boldsymbol{x}_{1},\boldsymbol{\theta}_{1} \right) \right)$ to the log-likelihood, and subjects with $z_{i}=0$ contribute

$$\begin{aligned} \log\left\{ \left( 1-\pi\left( \boldsymbol{x}_{1},\boldsymbol{\theta}_{1} \right) \right)\left[ F\left( R_{i};{\boldsymbol{x}_{2}\boldsymbol{,\theta}}_{2}, \lambda_{2},\beta\right)-F\left( L_{i};{\boldsymbol{x}_{2}\boldsymbol{,\theta}}_{2},\lambda_{2},\beta\right) \right] \right\}\#\left( 1 \right) \end{aligned}$$

to the log-likelihood as the difference of the cumulative densities is used to get the likelihood of disease developing between the left and right time points for interval-censored data. When $z_{i}$ is missing, it cannot be ruled out that the disease was present at baseline, so the left bound is fixed at zero and $F\left( L_{i};{\boldsymbol{x}_{2},\boldsymbol{\theta}}_{2},\lambda_{2},\beta\right)=0$. Therefore, subjects with missing $z_{i}$ contribute

$$\begin{aligned} \log\left\{ \pi\left( \boldsymbol{x}_{1},\boldsymbol{\theta}_{1} \right)+\left( 1-\pi\left( \boldsymbol{x}_{1},\boldsymbol{\theta}_{1} \right) \right)F\left( R_{i};{\boldsymbol{x}_{2}\boldsymbol{,\theta}}_{2},\lambda_{2},\beta\right) \right\}\#\left( 2 \right) \end{aligned}$$

to the log-likelihood. Putting these together, the complete-data log-likelihood is given by

$$\mathcal{l}_{c}\left( \boldsymbol{\theta} \right)=\sum_{z_{i}\text{observed}} \left[ z_{i}\log\left( \pi\left( \boldsymbol{x}_{1},\boldsymbol{\theta}_{1} \right) \right)+\left( 1-z_{i} \right)\left\{ \log\left( 1-\pi\left( \boldsymbol{x}_{1},\boldsymbol{\theta}_{1} \right) \right)+\log\left( F\left( R_{i};{\boldsymbol{x}_{2}\boldsymbol{,\theta}}_{2},\lambda_{2},\beta\right)-F\left( L_{i};{\boldsymbol{x}_{2}\boldsymbol{,\theta}}_{2},\lambda_{2},\beta\right) \right) \right\} \right]$$

$$\begin{aligned} + \sum_{z_{i} \text{missing}} \log\left( \pi\left( \boldsymbol{x}_{1},\boldsymbol{\theta}_{1} \right)+\left( 1-\pi\left( \boldsymbol{x}_{1},\boldsymbol{\theta}_{1} \right) \right)F\left( R_{i};{\boldsymbol{x}_{2}\boldsymbol{,\theta}}_{2},\lambda_{2},\beta\right) \right) \left( 3 \right) \end{aligned}$$

where $\boldsymbol{\theta}=\left\{ \boldsymbol{\theta}_{1}\boldsymbol{,}\boldsymbol{\theta}_{2},\lambda_{2},\beta\right\}$.

**1.2 Expectation step**

Given the current parameter estimates $\boldsymbol{\theta}^{\left( l \right)}$ and the observed data, the expected value of the latent variable $z_{i}$, $\mathbb{E}\left( z_{i}|\boldsymbol{\theta}^{\left( l \right)} \right)$, is estimated. When $z_{i}$ is known then the expected value of $z_{i}$ is simply its value. When $z_{i}$ is unknown, the left bound is always zero, so $F\left( L_{i};{\boldsymbol{x}_{2}\boldsymbol{,\theta}}_{2},\lambda_{2},\beta\right)=0$, and the expected value of $z_{i}$ is given by

$$\begin{aligned} \mathbb{E}\left( z_{i}|\boldsymbol{\theta}^{\left( l \right)} \right)=\frac{\pi\left( \boldsymbol{x}_{1},\boldsymbol{\theta}_{1}^{\left( l \right)} \right)}{\pi\left( \boldsymbol{x}_{1},\boldsymbol{\theta}_{1}^{\left( l \right)} \right)+\left( 1-\pi\left( \boldsymbol{x}_{1},\boldsymbol{\theta}_{1}^{\left( l \right)} \right) \right)F\left( R_{i};{\boldsymbol{x}_{2}\boldsymbol{,}\boldsymbol{\theta}_{2}^{\left( l \right)},\lambda}_{2}^{\left( l \right)},\beta^{\left( l \right)} \right)}. \#\left( 4 \right) \end{aligned}$$

**1.3 Maximization step**

Given the observed data and the latent variables, we estimate the parameters by maximizing the expected complete data log-likelihood, $Q\left( \boldsymbol{\theta}|\boldsymbol{\theta}^{\left( l \right)} \right)$

$$\begin{aligned} Q\left( \boldsymbol{\theta} | \boldsymbol{\theta}^{\left( l \right)} \right)= \sum_{i=1}^{n} \mathbb{E}\left( z_{i} | \boldsymbol{\theta}^{\left( l \right)} \right)\log\left( \pi\left( \boldsymbol{x}_{1},\boldsymbol{\theta}_{1} \right) \right)+ \end{aligned}\left( 1\mathbb{-E}\left( z_{i}|\boldsymbol{\theta}^{\left( l \right)} \right) \right)\left[ \log\left( 1-\pi\left( \boldsymbol{x}_{1},\boldsymbol{\theta}_{1} \right) \right) \right.$$

$$\left. + log \left( F\left( R_{i};{\boldsymbol{x}_{2}\boldsymbol{,\theta}}_{2},\lambda_{2},\beta\right)-F\left( L_{i};{\boldsymbol{x}_{2}\boldsymbol{,\theta}}_{2},\lambda_{2},\beta\right) \right) \right]. (5)$$

To update the parameters, we use a single Newton step where the next parameter value is given by

$$\begin{aligned} \boldsymbol{\theta}^{\left( l+1 \right)}=\boldsymbol{\theta}^{\left( l \right)}-\left[ \text{H}_{Q}\left( \boldsymbol{\theta}^{\left( l \right)} \right) \right]^{-1}\text{U}_{Q}\left( \boldsymbol{\theta}^{\left( l \right)} \right). \#\left( 6 \right) \end{aligned}$$

In the equation above, $\text{U}_{Q}\left( \boldsymbol{\theta} \right)$ is the gradient of $Q\left( \boldsymbol{\theta}|\boldsymbol{\theta}^{\left( l \right)} \right)$ consisting of first derivatives, and $\text{H}_{Q}\left( \boldsymbol{\theta} \right)$ is the Hessian matrix of second partial derivatives of $Q\left( \boldsymbol{\theta}|\boldsymbol{\theta}^{\left( l \right)} \right)$.

The expectation and maximisation step are repeated until the absolute change in the log-likelihood between iterations is less than ${10}^{-8}$. After the model has converged, the Hessian of the observed log-likelihood is calculated using numerical differentiation, then variances of the parameter estimates are calculated by the inverse of the negative of the Hessian of the observed data log-likelihood at $\hat{\boldsymbol{\theta}}$.

**1.4 Initial values**

Direct implementation of (3) and (4) will lead to convergence problems. Instead,  a two-step process is implemented in order to estimate the background risk. To find initial values for the other model parameters, we use a version of the Short EM algorithm^19^. This method involves using multiple sets of random starting values and performing short runs of the EM algorithm with a relaxed convergence criterion (e.g., ${10}^{-2}$ instead of ${10}^{-8}$), a low number of maximum iterations (e.g., 10) and background risk fixed at zero. The parameter estimates that yield the highest log-likelihood are then used as initial values in the EM gradient algorithm.

In the second step, we take a very small value of background risk (e.g., fixed at $\log\left( \beta\right)= -12$) with the initial values found in the previous step for the remaining parameters, then we iteratively increase the fixed value of background risk and at each step use as initial values for the remaining parameters the estimates found in the previous iteration until the log-likelihood stops increasing. Lastly, the estimates from the most recent iteration are used as initial values in a final run of the EM algorithm where the background risk is no longer kept at a fixed value to find maximum likelihood estimates with standard errors for all parameters. This procedure yields a high-speed algorithm with a high convergence rate which makes it suitable for general consumption; speed and convergence results are described in the simulation study.
